# Ancestry-specific TWAS refines type 2 diabetes GWAS loci in disease-relevant tissues

**DOI:** 10.64898/2026.08.19.26360695

**Authors:** Inti Pagnuco, Stephen Eyre, Magnus Rattray, Andrew P Morris

## Abstract

Type 2 diabetes (T2D) is a complex metabolic disorder characterized by hyperglycemia and insulin resistance. Although genome-wide association studies (GWAS) have identified >600 T2D risk loci, the causal genes and the relevant tissues mediating these associations remain largely unresolved.

To address this challenge, we performed tissue-specific, ancestry-aware transcriptome-wide association studies (TWAS) across six T2D-relevant tissues: subcutaneous adipose, visceral adipose, brain hypothalamus, liver, skeletal muscle, and pancreas. We conducted ancestry-specific multi-tissue TWAS in European ancestry (EUR) data using summary statistics from the largest EUR GWAS (242,283 cases and 1,569,734 controls) and pre-trained gene expression prediction models derived from 689 EUR individuals from the Genotype-Tissue Expression (GTEx) Project. Conditional analyses were performed to identify independent TWAS signals.

We identified 684-750 significant gene-T2D associations per tissue (P < 1.919 × 10^−6^), implicating both established and novel candidate genes. Among these, *JAZF1* and *IDE* showed consistent association signals across all six tissues, whereas *TCF7L2* and *WSF1* exhibited heterogeneous effects restricted to a subset of T2D-relevant tissues. Conditional analyses further refined these signals to 289–322 independent TWAS signals per tissue. Together, these finding highlight substantial regulatory heterogeneity in the genetic architecture of T2D and underscore the importance of tissue context in interpreting disease-associated loci.

Cross-ancestry replication of EUR-derived TWAS signals was evaluated in African American (AFA) individuals. We conducted an AFA-TWAS using summary statistics from the largest AFA GWAS (50,251 cases and 103,909 controls) in combination with gene expression prediction models trained in 111 AFA individuals from GTEx. We observed significant enrichment of EUR-derived T2D TWAS signals in the AFA TWAS across subcutaneous adipose, visceral adipose, skeletal muscle, and pancreas, whilst enrichment was weaker in liver, likely reflecting limited sample size.

Overall, our findings demonstrate that integrating tissue-specific and ancestry-aware TWAS refines the identification of causal genes for T2D, with cross-ancestry replication supporting the robustness of these signals and cross-tissue analyses revealing context-specific effects. However, they also highlight the limited availability of non-EUR datasets and the need for larger, more diverse ancestry-specific transcriptomic resources.

## 1. Introduction

Type 2 diabetes (T2D) is a complex metabolic disorder characterized by chronic hyperglycemia resulting from a combination of insulin resistance and impaired pancreatic β-cell function. The regulation of glucose homeostasis depends on the coordinated activity of multiple metabolic tissues that control insulin secretion, glucose uptake, and systemic metabolic balance (Daryabor et al., 2020; DeFronzo et al., 2015; Galicia-Garcia et al., 2020). Due to its high and increasing prevalence worldwide, T2D represents a major global public health challenge (DeFronzo et al., 2015).

Genome-wide association studies (GWAS) have advanced our understanding of the genetic architecture of T2D, with large-scale meta-analyses identifying >600 susceptibility loci (Mahajan et al., 2018; Suzuki et al., 2024; Vujkovic et al., 2020). However, most GWAS signals map to non-coding regions, making it difficult to interpret their biological impact. One prevailing explanation is that these variants influence disease risk primarily through regulation of gene expression rather than altering protein-coding sequence (Maurano et al., 2012). Interpreting GWAS findings therefore requires not only the identification of causal genes but also an understanding of the tissues in which they act, since gene regulation is highly tissue-specific and the same variant can have distinct effects across different tissues (Fu et al., 2012). This consideration is particularly relevant for T2D, which involves biological processes across multiple metabolic tissues, including pancreatic islets, liver, adipose, brain, and skeletal muscle.

Transcriptome-wide association studies (TWAS) have emerged as powerful method to link genetic variants to gene expression and provide a functional interpretation of GWAS loci (Gamazon et al., 2015; Gusev et al., 2016; Hu et al., 2019). TWAS uses gene expression prediction models, which capture the effects of expression quantitative trait loci (eQTLs), to impute genetically regulated expression levels in independent cohorts using only genotype data. By integrating these models with GWAS summary statistics, TWAS identifies genes whose predicted expression is associated with complex traits. This approach not only helps prioritize candidate genes at GWAS loci but also provides insights into the tissues through which genetic variants influence disease risk.

A key limitation of both GWAS and TWAS is the underrepresentation of diverse populations. To date, most studies and gene expression reference panels have been generated primarily from European ancestry (EUR) individuals, which limits the generalizability of findings to other population groups. This limitation arises in part because differences in allele frequencies and linkage disequilibrium patterns across ancestry groups can affect the detection of genetic associations and reduce the performance of gene expression prediction models in non-EUR populations (Adeyemo & Rotimi, 2010; Carlson et al., 2013; Pagnuco et al., 2025). Expanding representation across ancestries is therefore essential to improve locus discovery, refine fine-mapping resolution, and enable more accurate and equitable interpretation of genetic risk across populations (Vujkovic et al., 2020).

To overcome these challenges, ancestry-aware and cross-tissue approaches have been developed. Resources such as the Ancestry-specific cross-tissue Gene Expression Model database (AGEMdb) leverage cross-tissue modelling to enable gene expression prediction across multiple tissues for EUR and African American (AFA) studies (Pagnuco et al., 2026). AGEMdb was built using the Unified Test for Molecular SignaTures (UTMOST) framework (Hu et al., 2019), which is one of several existing methods that jointly model gene expression across tissues (Barbeira et al., 2019; Zhou et al., 2020). These approaches are motivated by the observation that genetic regulation is often shared across tissues(The GTEx Consortium et al., 2015), allowing correlated expression patterns to improve prediction performance and robustness (Pagnuco et al., 2025; Zhou et al., 2020). This integration of ancestry-aware and cross-tissue approaches enables more accurate and transferable TWAS analyses across ancestry groups (Pagnuco et al., 2025).

In this study, we apply a tissue-specific, ancestry-aware TWAS framework to refine T2D GWAS signals. By integrating GWAS summary statistics with gene expression prediction models across relevant metabolic tissues, we aim to identify candidate genes underlying T2D risk loci and infer their likely tissues of action. TWAS signals are further refined through conditional analyses, and their cross-ancestry generalizability is assessed. Collectively, this approach enhances the functional interpretation of GWAS findings, provides deeper insight into the molecular mechanisms underlying T2D susceptibility, and extends these insights to underrepresented populations.

## 2. Methods

### 2.1. Gene Expression Imputation Models for TWAS in T2D-relevant tissues

To perform the TWAS, we downloaded cross-tissue gene expression imputation models from the Ancestry-specific Gene Expression Model Database (AGEMdb) (Pagnuco et al., 2026). These ancestry-specific models were trained using 689 EUR and 111 AFA participants from the Genotype-Tissue Expression (GTEx) Project (The GTEx Consortium et al., 2015) and constructed using the Cross-Tissue gene expression IMPutation (CTIMP) framework implemented in UTMOST (Hu et al., 2019).

For the EUR dataset, gene expression imputation models were available for six T2D-relevant tissues: subcutaneous adipose, visceral omentum adipose, brain hypothalamus, liver, skeletal muscle, and pancreas. Each model was trained with between 134 and 514 samples per tissue. For the AFA dataset, gene expression imputation models were available for only five tissues because only 10 samples were available for brain hypothalamus. For the remaining five tissues, each model was trained with between 17 and 77 samples. Supplementary Table 1 provides the detailed sample size per tissue, as well as the total number of gene models available in AGEMdb and the subset corresponding to protein-coding genes, which we focussed on for our downstream analyses.

### 2.2. T2D Discovery TWAS Analysis

Our discovery TWAS analysis was conducted in the EUR dataset across T2D-relevant tissues using the UTMOST framework (Hu et al., 2019). Analyses integrated the EUR gene expression imputation models with summary statistics from the largest available EUR T2D GWAS (Suzuki et al. 2024), comprising 242,283 cases and 1,569,734 controls. Briefly, UTMOST predicts genetically regulated gene expression across multiple tissues using eQTL-based imputation models and subsequently tests the association between predicted expression and T2D risk within each tissue.

To account for multiple testing, a Bonferroni correction was applied based on the total number of genes tested in the TWAS across all T2D-relevant tissues (26,048 genes; Supplementary Table 1), yielding a transcriptome-wide significance threshold of *P* < 1.919 × 10^−6.^ Subsequent analyses were restricted to protein-coding genes to facilitate biological interpretation.

### 2.3 Refinement of T2D TWAS signals

To refine TWAS signals and identify independent gene-level associations, we performed stepwise conditional analyses using UTMOST (Hu et al., 2019). Genomic regions were defined by first selecting protein-coding genes demonstrating transcriptome-wide significant associations with T2D (P < 1.919 × 10^−6^). For each significant gene, we constructed a genomic window extending 1 Mb upstream of the transcription start site and 1 Mb downstream of the transcription stop site, corresponding to the boundaries for SNPs used for training the gene expression imputation models. When windows from multiple significant genes overlapped, they were merged into a single multi-gene region. Within each multi-gene region, we identified the “lead gene” with the strongest TWAS signal and conditioned the association of other genes in the region on its predicted expression. Any gene-level association that remained transcriptome-wide significant after conditioning was considered independent of the lead gene in the region. In cases where conditional analyses produced unstable results, genes contributing to the instability were removed and the analysis was repeated until a stable set of independent transcriptome-wide significant genes was obtained. This procedure was repeated until no genes met the transcriptome-wide significance threshold after conditioning and the TWAS signals across the multi-gene region were fully explained.

### 2.4. Cross-ancestry validation of T2D TWAS signals

To assess cross-ancestry transferability of transcriptome-wide significant EUR-T2D TWAS signals, we applied UTMOST using AFA tissue-specific gene expression models and summary statistics from the largest available AFA T2D GWAS (Suzuki et al., 2024) comprising 50,251 cases and 103,909 controls.

First, all transcriptome-wide significant protein coding genes identified in the EUR-TWAS analysis were collected for the five T2D-relevant tissues available in the AFA dataset. Among these, we determined how many had corresponding gene expression imputation models available in the AFA dataset for the same tissue. For these genes, replication was evaluated in two ways: (i) by assessing enrichment of genes with nominal significance in AFA TWAS (P < 0.05); and (ii) by assessing enrichment of genes with the same direction of effect in AFA and EUR datasets. One-sided binomial tests were used to determine whether the observed proportions of nominally validated genes and concordant effect direction genes were higher than expected by chance.

## 3. Results

### 3.1. Gene-level T2D associations in EUR TWAS

The total number of gene models available for T2D-relevant tissues from AGEMdb ranged from 23,277 to 32,644 per tissue in the EUR dataset (Supplementary Table 1). UTMOST association analyses between genetically imputed gene expression and T2D were conducted using the EUR GWAS summary statistics for between 18,092 and 21,704 genes, depending on the tissue (Table 1). The reduction in the number of genes tested reflects incomplete SNP coverage in the GWAS required to support tissue-specific gene expression imputation models. Protein-coding gene-level associations reaching transcriptome-wide significance (P < 1.919 × 10^−6^) are detailed in Supplementary Table 2, with the number of significant genes per tissue ranging from 684 in skeletal muscle to 750 in subcutaneous adipose (Table 1). Figure 1 presents Manhattan plots of T2D TWAS signals for each tissue and all tested protein-coding genes.

**Figure 1.**
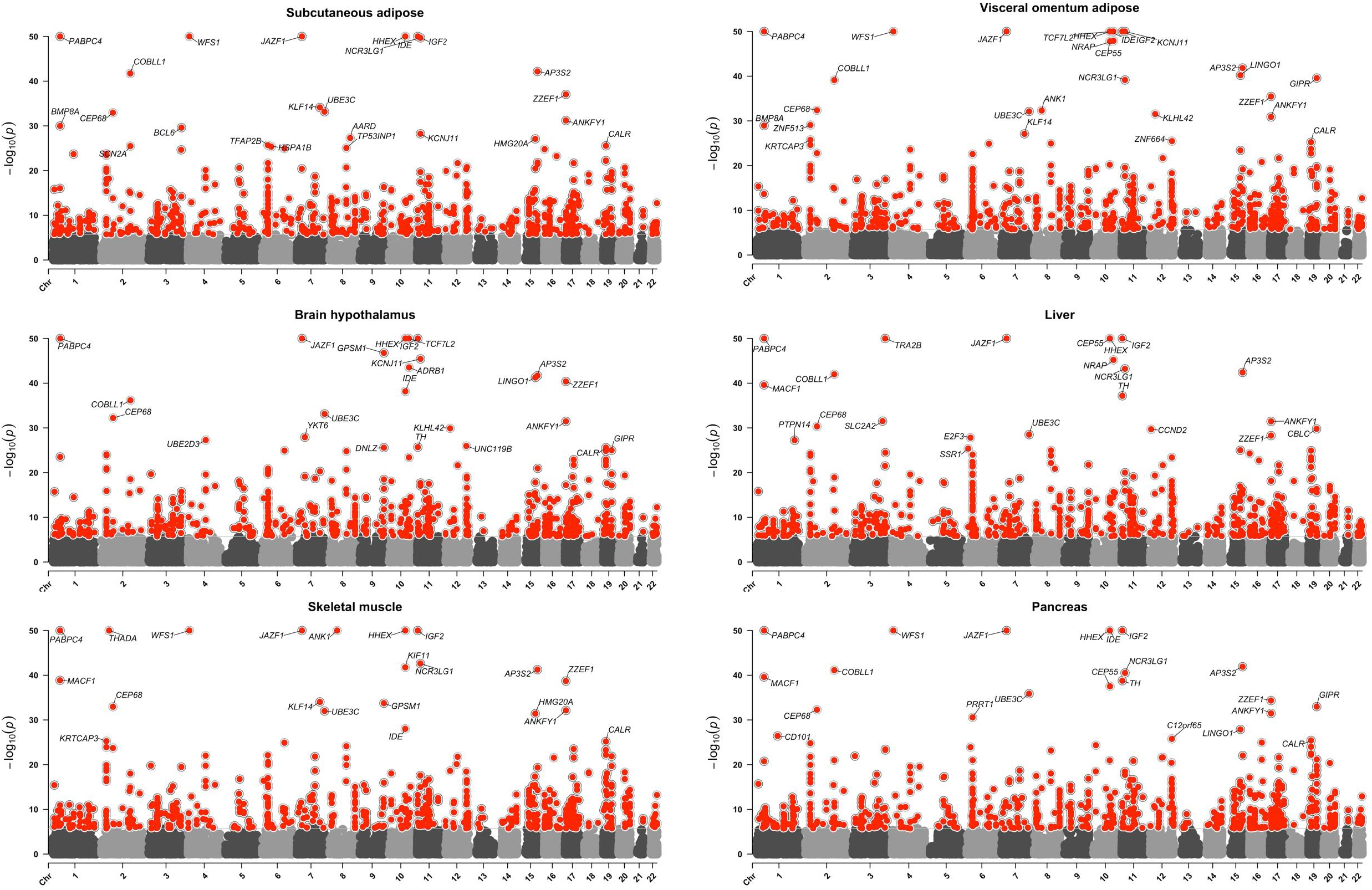
Manhattan plots of TWAS associations for protein-coding genes across T2D-relevant tissues. Each plot shows ™log10(P) values for all tested protein-coding genes, with transcriptome-wide significant genes (P < 1.919 × 10^−6^) highlighted in red. For visualization, P-values smaller than 1e-50 were capped.

We considered all protein-coding genes that achieved transcriptome-wide significance in at least one tissue (Supplementary Table 3). Among the 1,448 genes identified, 1,268 mapped to loci previously associated with T2D, as reported by Suzuki et al., (2024), while 180 represent novel associations. Examples of these novel genes include *CEP55* (significant in adipose subcutaneous, p = 2.82 × 10^−8^; visceral adipose, p = 1.59 × 10^−48^; liver, p = 1.70 × 10^−79^; pancreas, p = 2.96 × 10^−38^), *EXT1* (significant in brain hypothalamus, p = 1.91 × 10^−9^; liver, p = 1.40 × 10^−21^; pancreas, p = 8.74 × 10^−7^), and *HOXA13* (significant in adipose subcutaneous, p = 3.95 × 10^−21^), with other tested tissues either not reaching TWAS significance or lacking an available prediction model.

To evaluate cross-tissue association patterns for T2D, we focused on the subset of 1,264 genes (of 1,448 total) with predictive models available across all six tissues. Of these genes, 425 (34%) attained transcriptome-wide significance in only one tissue, whilst 277 (22%) reached transcriptome-wide significance in all six tissues. Of these 277 genes with significant T2D associations across all tissues, 155 genes (56%) showed the same direction of effect in every tissue. These results indicate that, while most genes have broadly shared regulatory effects on T2D, a substantial fraction exhibit tissue-specific or context-dependent influences.

The strongest overall T2D association was observed for *JAZF1*, which showed highly significant and consistently negative effects (increased expression associated with decreased T2D risk) across all six tissues (subcutaneous adipose: β = -0.432, p = 2.80 × 10^−91^; visceral omentum adipose: β = -0.395, p = 1.70 × 10^−91^; brain hypothalamus: β = -1.991, p = 1.12 × 10^−80^; liver: β = -0.582, p = 8.27 × 10^−91^; skeletal muscle: β = -0.366, p = 1.73 × 10^−91^; pancreas: β = -0.291, p = 1.61 × 10^−91^), consistent with a broadly shared regulatory role. *JAZF1* is a transcriptional cofactor that plays a functional role in glucose regulation, lipid homeostasis and inflammation (Liao et al., 2019). In contrast, *TCF7L2* exhibited a tissue-specific association pattern, with significant effects in different directions on T2D risk in subcutaneous adipose (β = 0.301, p = 1.32 × 10^−9^), visceral adipose (β = -0.965, p = 7.68 × 10^−85^), and brain (β = 0.951, p = 9.19 × 10^−82^). *TCF7L2* is a transcription factor in the Wnt signalling pathway that has essential developmental and metabolic roles in adipose tissue and functions by regulating expression of genes involved in glucose and lipid metabolism (Geoghegan et al., 2019). Finally, *RCN2* represents the strongest tissue-specific T2D association signal, being significant only in liver (β = 0.301, p = 1.00 × 10^−25^), indicating that liver is the primary tissue driving regulatory effects underlying this association. RCN2 has an established role in regulation of lipid metabolism and lipolysis, and elevated RCN2 activity impacts on hepatic liver handling.

### 3.2. Refinement of T2D TWAS signals

Using the protein-coding genes identified as significant in the EUR-TWAS analysis, we performed conditional analysis to identify independent gene-level association signals, excluding genes located within the Major Histocompatibility Complex (MHC). After conditioning, the number of independent signals at transcriptome-wide significance ranged from 289 in liver to 322 in subcutaneous adipose (Table 1). Detailed results of the conditional analyses are provided in Supplementary Tables 4-5. For many multi-gene regions, the number of independent TWAS signals after conditional analysis was reduced to just one or two gene-level associations per tissue, reflecting shared underlying regulatory SNPs (or linkage disequilibrium between them). For example, in a 6.4 Mb region of chromosome 8 (7.3–13.7 Mb), multiple protein-coding genes showed transcriptome-wide significant associations across all examined tissues, ranging from 9 to 14 genes per tissue. The lead gene signals were *PPP1R3B* in subcutaneous adipose (p = 1.14 × 10^−14^), *PINX1* in visceral omentum adipose (p = 1.04 × 10^−13^) and liver (p = 1.04 × 10^−13^), *MSRA* in skeletal muscle (p = 6.98 × 10^−14^), and *XKR6* in brain (p = 6.28 × 10^−19^) and pancreas (p = 3.43 × 10^−18^). After conditional analysis, only the tissue-specific lead gene remained as an independent signal in most tissues, fully accounting for the gene-level association at this locus. This pattern was observed for *PINX1* in visceral omentum adipose and liver, *MSRA* in skeletal muscle, and *XKR6* in pancreas and brain hypothalamus. In contrast, subcutaneous adipose tissue retained two independent signals, *PPP1R3B* (p= 3.52 × 10^−14^) and *LONRF1* (p= 3.32 × 10^−8^), highlighting a more complex association pattern in this tissue.

A similar pattern of refinement was observed across a 4.6 Mb region of chromosome 16 (27.3– 31.9 Mb), where 16 to 26 protein-coding genes per tissue were initially associated with T2D at transcriptome-wide significance. The lead association signals corresponded to *TMEM219* in subcutaneous adipose (p = 3.36 × 10^−14^), brain (p = 7.17 × 10^−15^), and pancreas (p = 7.17 × 10^−14^), and to *INO80E* in visceral omentum adipose (p = 2.20 × 10^−14^), skeletal muscle (p = 4.33 × 10^−15^), and liver (p = 5.06 × 10^−13)^. Conditional analysis refined the locus to a single independent gene per tissue, indicating that the association was driven by the respective lead gene in most tissues: *TMEM219* in subcutaneous adipose, pancreas, and brain hypothalamus, and *INO80E* in liver and skeletal muscle. Visceral omentum adipose was the only tissue retaining two independent signals (*INO80E*, p = 2.6 × 10^−11^ and *IL27* p = 1.31 × 10^−6^) suggesting partially distinct regulatory mechanisms within this locus.

Together, these examples demonstrate that conditional analysis substantially refines broad multi-gene association regions into a small number of independent signals and reveals tissue-specific differences of genes driving the observed T2D associations.

### 3.3. Cross-ancestry validation of TWAS signals

To evaluate cross-ancestry transferability of TWAS signals, we applied UTMOST to test AFA gene expression imputation models using AFA T2D GWAS summary statistics (Suzuki et al., 2024) for subcutaneous adipose, visceral adipose, liver, skeletal muscle and pancreas. We then assessed enrichment of EUR TWAS gene associations at transcriptome-wide significance in the AFA dataset across these T2D-relevant tissues (Figure 2 and Supplementary Table 6). Among EUR-significant protein-coding genes with available AFA predictive models, we observed significant enrichment for nominal replication (P < 0.05) in subcutaneous adipose (86/688, p_binomial_ = 1.36 × 10^-14^), visceral omentum adipose (77/666, p_binomial_ = 1.50 × 10^-11^), skeletal muscle (78/638, p_binomial_ = 6.84 × 10^-13^), and pancreas (72/675, p_binomial_ = 2.26 × 10^-09^), but not in liver (18/256, p_binomial_ = 9.33 × 10^-2^). Enrichment for concordance of effect direction between EUR and AFA TWAS was also significant in subcutaneous adipose (419/688, p_binomial_ = 5.90 × 10^-09^), visceral omentum adipose (379/666, p_binomial_ = 2.06 × 10^-04^), skeletal muscle (409/638, p_binomial_ =4.89 × 10^-13^), and pancreas (386/675, p_binomial_ =1.07 × 10^-04^), but not in liver (133/256, p_binomial_ = 0.28). These results support tissue-specific, cross-ancestry replication of TWAS associations, with the lack of replication in liver likely reflecting the smaller sample size of this tissue in the AFA dataset.

**Figure 2.**
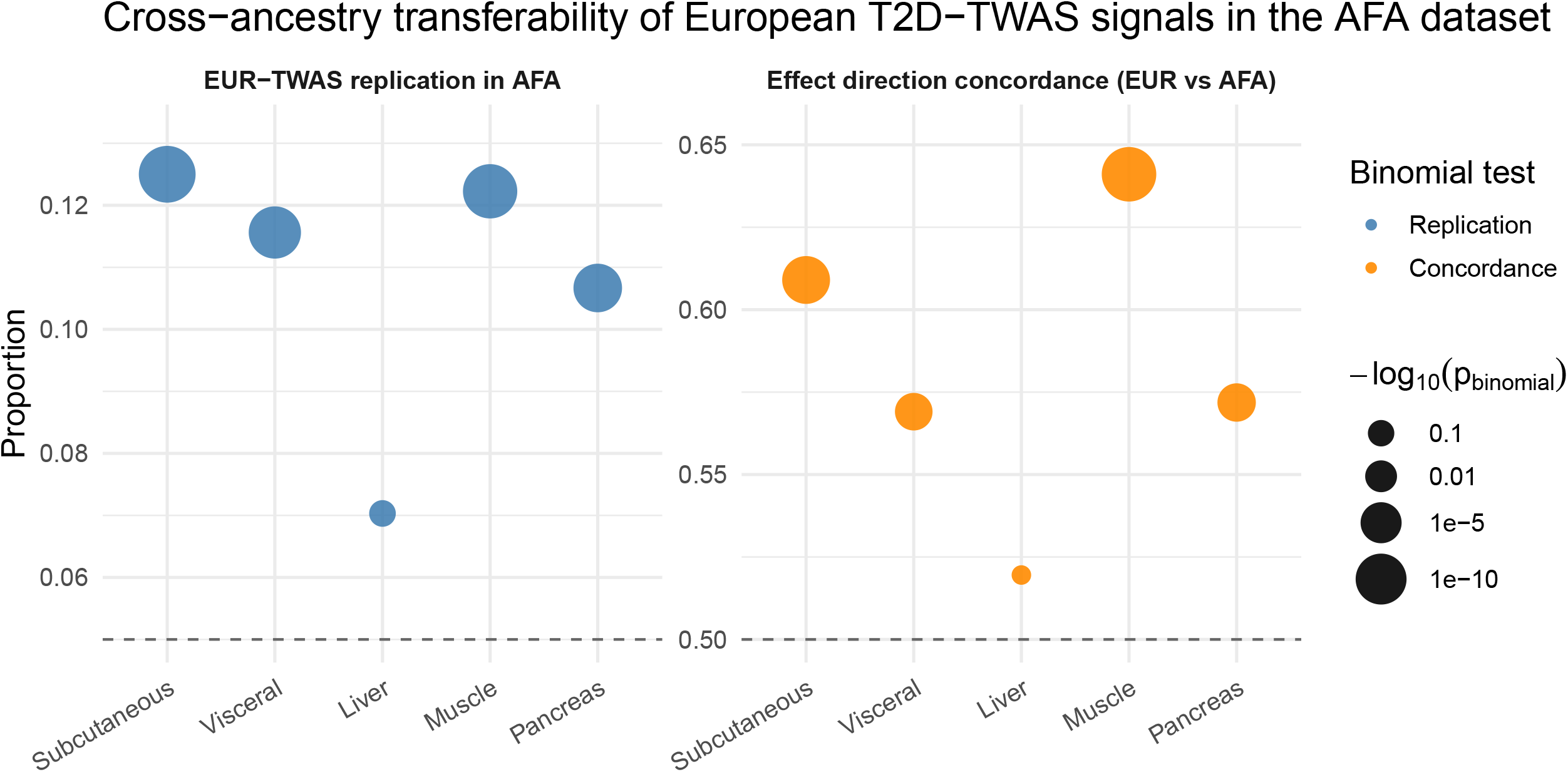
Cross-ancestry transferability of European T2D-TWAS signals in African American ancestry cohort across tissues. Replication and effect direction concordance of TWAS associations identified in European (EUR) ancestry data were evaluated in African ancestry (AFA) datasets across five T2D-metabolically relevant tissues. Left panel shows the proportion of EUR-significant genes (with available AFA prediction models) that nominally replicated in AFA TWAS analyses (P < 0.05). Right panel shows the proportion of genes with concordant effect direction between EUR and AFA TWAS associations. Each point represents a tissue. Point size reflects statistical significance of enrichment (™log10 binomial test P-value). Dashed horizontal lines indicate the nominal replication threshold (P = 0.05) and random expectation for concordance (0.5). Significant enrichment was observed in subcutaneous adipose, visceral omentum adipose, skeletal muscle, and pancreas, but not liver.

## 4. Discussion

In this study, we implemented a tissue-specific and ancestry-aware TWAS framework across six T2D-relevant tissues to improve understanding of the causal genes and tissue-specific regulatory mechanisms driving T2D susceptibility. By integrating EUR-based T2D-TWAS findings with conditional fine-mapping and further evaluating cross-ancestry transferability using AFA models, we identified robust gene-trait associations that are biologically relevant across populations. Our results provide a more refined understanding of the tissue-specific contexts underlying T2D genetic risk, helping to bridge the gap between GWAS loci and functional mechanisms, and underscoring the importance of population diversity in cross-ancestry TWAS replication. Our findings highlight the value of evaluating T2D-associated genes across multiple disease-relevant tissues. This is consistent with the rationale underlying cross-tissue TWAS frameworks such as UTMOST, which leverage shared regulatory eQTL effects across tissues to improve expression prediction and gene discovery (Hu et al., 2019). While several genes in our study showed consistent associations across tissues, others exhibited marked heterogeneity in effect size and direction, demonstrating that multi-tissue analyses can reveal both shared and tissue-specific regulatory mechanisms underlying T2D susceptibility.

The value of this multi-tissue resolution is illustrated by comparison with previous TWAS studies focused on individual tissues. Tang et al., 2023 used an integrative framework combining TWAS, Mendelian randomisation, colocalisation, and functional analyses, to prioritise five putative causal T2D genes in visceral adipose tissue. All five genes (*PABPC4, CWF19L1, CCDC92, CCNE2*, and *HAUS6*) were also significant across all T2D-relevant tissues in our analysis. *PABPC4, CWF19L1*, and *CCDC92* showed consistently negative effects, while *CCNE2* and *HAUS6* exhibited more heterogeneous patterns across tissues, suggesting context-dependent regulation. These results extend adipose-specific findings by showing that these genes have effects that are broadly present across T2D-relevant tissues, with tissue-specific differences in effect strength and direction.

Beyond characterising the tissue specificity of individual genes, multi-tissue TWAS approaches can also improve interpretation of complex association signals at individual T2D risk loci. TWAS associations frequently involve multiple genes at the same locus, and conditional analysis can disentangle these signals into independent, tissue-specific associations, thereby clarifying the biological contexts through which genes influence disease risk. In our study, UTMOST conditional analyses helped distinguish independent associations across tissues, highlighting the value of incorporating tissue context into gene prioritisation. More broadly, emerging TWAS methodologies continue to increase biological resolution. For example, scPrediXcan (Y. Zhou et al., 2025), integrates single-cell transcriptomic data and deep learning approaches to identify cell-type-specific gene–trait associations in complex diseases, including T2D. Together, these strategies demonstrate that integrating complementary analytical frameworks can refine TWAS signals into more biologically interpretable, context-specific associations and improve prioritisation of candidate causal genes.

Our cross-ancestry analyses demonstrate that a substantial proportion of EUR-derived T2D TWAS associations are transferable to AFA-derived TWAS signals, with significant enrichment of associations and concordant effect directions across subcutaneous and visceral adipose tissue, skeletal muscle, and pancreas. These findings suggest that the effects of genetically regulated gene expression are largely conserved across ancestries, despite known differences in linkage disequilibrium structure, allele frequencies, and eQTL architecture (Mogil et al., 2018). This observation is consistent with a large multi-ancestry T2D-GWAS, which demonstrated substantial sharing of the genetic architecture underlying T2D across populations and showed that increased ancestral diversity improves fine-mapping resolution and the localisation of putative causal variants (Mahajan et al., 2022). The absence of significant enrichment in liver likely reflects the limited availability of transcriptomic data from AFA individuals, resulting in reduced statistical power, rather than a genuine lack of shared biological mechanisms underlying T2D susceptibility.

Our study has several limitations that are inherent to the TWAS framework. TWAS relies on genetically predicted gene expression models, which are primarily constructed from common cis-eQTLs. As such, they capture only the heritable component of gene expression explained by cis-acting variants and do not account for rare cis variants, trans-regulatory effects, environmental influences, or post-transcriptional regulatory mechanisms. Notably, common cis-eQTLs explain only a modest proportion of the total genetic variance in gene expression (∼9–12%), which limits the overall explanatory power of these models (Grundberg et al., 2012). Model performance also varies across tissues, depending on tissue-specific eQTL architecture and the size of reference transcriptomic datasets, and across populations, due to ancestry-specific differences in genetic architecture. Incomplete SNP coverage in GWAS further restricts the number of genes that can be evaluated, potentially leading to underestimation of true associations.

Despite these limitations, the study has several notable strengths. A major strength is the integration of tissue-specific TWAS, conditional fine-mapping, and cross-ancestry replication within a unified analytical framework. The use of ancestry-specific gene expression models from AGEMdb enabled a systematic evaluation of T2D-associated genes across multiple disease-relevant tissues and ancestries, while conditional fine-mapping allowed identification of independent gene-level associations. Importantly, the cross-ancestry analyses provided evidence that many genetically regulated expression signals are shared across ancestries, highlighting the value of incorporating diverse populations in genomic studies of complex traits. Expanding this work will require larger genotypic and transcriptomic datasets from multiple populations, which would further improve gene discovery and enable more comprehensive investigation of allelic heterogeneity and population-specific eQTLs.

In conclusion, our findings demonstrate that integrating tissue-specific and ancestry-aware TWAS refines the identification of candidate causal genes for T2D, with cross-ancestry replication providing support for the robustness of these associations and cross-tissue analyses revealing context-specific effects. These results advance our understanding of the genetic architecture and biological mechanisms underlying T2D and highlight promising targets for future functional investigation and therapeutic development. At the same time, they underscore the limited availability of non-EUR transcriptomic resources and the need for larger, more diverse ancestry-specific datasets to improve gene discovery and ensure the equitable translation of genomic research across populations.

## Data Availability

All data produced in the present study are available upon reasonable request to the authors

## Figures and Tables legend

**Table 1**. Summary of EUR-TWAS results across six T2D-relevant tissues. For each tissue, the table includes: the number of genes tested in the EUR-TWAS analysis (genes with both an expression prediction model and corresponding GWAS data), the number of protein-coding genes reaching transcriptome-wide significance (Bonferroni-corrected P < 1.919 × 10^−6^), the number of TWAS-significant genes included in conditional analysis (excluding genes with genomic regions overlapping the Major Histocompatibility Complex (MHC)), and the total number of independent signals identified following stepwise conditional analysis.

## Supplementary tables

**Supplementary Table 1**. GTEx sample sizes and gene expression prediction models from AGEMdb (Ancestry-specific Gene Expression Model Database) by tissue and ancestry group. For each tissue, the table displays the number of GTEx samples used to train gene expression prediction models, the total number of available gene expression models in AGEMdb, and the number of protein-coding gene models—representing a subset of the total models—separated for European (EUR) and African American (AFA) ancestries.

**Supplementary Table 2**. Significant protein-coding genes identified by EUR-TWAS across six tissues relevant to T2D. Transcriptome-wide association (TWAS) results for European-ancestry individuals across six Type 2 diabetes relevant tissues: subcutaneous adipose, visceral adipose (omentum), brain hypothalamus, liver, skeletal muscle, and pancreas. Genes are ordered according to their genomic coordinates, with results from all tissues aligned side by side. The table includes all protein-coding genes that reached significance in at least one tissue. Significant gene–tissue associations are displayed in black text, whereas non-significant associations are shown in light grey text. A dash (“–”) indicates that the gene was either not evaluated in that tissue or that no result was available due to insufficient GWAS coverage.

For each tissue, the table reports the Ensembl gene identifier (Ensembl ID), gene symbol (gene_name) and genomic coordinate, together with the TWAS association P-value (pvalue, P) and estimated effect size (effect_size, β). Additional metrics include the predicted expression variance (var_g); the number of SNPs tested in the TWAS model (Number of SNPs tested), which depends on the availability of SNPs in the corresponding GWAS summary statistics; the number of SNPs used to estimate the covariance structure (Number of covariance SNPs); and the total number of SNPs included in the gene expression prediction model (Number of model SNPs).

TWAS analyses were performed using European ancestry-specific gene expression prediction models from AGEMdb and European ancestry T2D GWAS summary statistics within the UTMOST framework.

**Supplementary Table 3**. EUR-TWAS association statistics for protein-coding genes significantly associated with T2D in at least tissue.

This table reports protein-coding genes that reached transcriptome-wide significance in at least one of six metabolically relevant tissues: subcutaneous adipose tissue, visceral omental adipose tissue, hypothalamus, liver, skeletal muscle, and pancreas. For each gene, the Ensembl gene ID, gene symbol, and genomic coordinates are provided, together with the effect direction pattern across tissues (based on the sign of β), the number of tissues in which the gene reached statistical significance, and the minimum P-value observed across tissues. Tissue-specific TWAS association P-values and corresponding effect sizes (β) are reported for each tissue. A dash (“–”) indicates that the gene was not tested in the corresponding tissue because no gene expression prediction model was available or due to insufficient GWAS coverage. Significant associations are shown in black text, whereas non-significant associations and missing values (“–”) are shown in light grey. Only significant associations were included in the count of significant tissues. The effect direction pattern was determined using the sign of β across all tested tissues, excluding missing values (“–”). Genes are ranked according to their minimum P-value across tissues. The final column indicates whether each gene has previously been reported as associated with T2D by Suzuki et al. (2024).

**Supplementary Table 4**. Conditional P-values for independent gene-level associations with T2D across metabolically relevant tissues.

The table reports significant protein-coding genes identified as independent signals by conditional analysis across six metabolically relevant tissues: subcutaneous adipose, visceral omentum adipose, brain hypothalamus, liver, skeletal muscle, and pancreas. For each gene, the Ensembl ID and gene symbol are provided.

Each tissue column shows the P-value from the conditional analysis performed within that tissue, accounting for other independent signals at the same locus. When only one independent signal was identified in a tissue, conditional analysis was not applicable, and the reported value corresponds to the original (unconditional) association P-value. The “Minimum P-value across tissues” column indicates the smallest P-value observed for each gene across all tissues. A dash (“-”) indicates that the gene was not tested in that tissue, did not reach significance, or was not retained as an independent signal after conditional analysis. Genes are ranked according to their minimum P-value across tissues.

**Supplementary Table 5**. Independent gene signals identified after conditional analysis across T2D-relevant tissues. This table summarizes genomic regions (hg38 coordinates) in which conditional analyses were performed across six metabolically relevant tissues: subcutaneous adipose, visceral omentum adipose, brain hypothalamus, liver, skeletal muscle, and pancreas tissues.

For each tissue and genomic region, the table reports the genomic coordinates (hg38), the prioritized independent gene signal identified after the conditional analysis (Ensemble ID and gene symbol), the primary EUR-TWAS association P-value, and the conditional P-value obtained after adjustment for other independent signals at the same locus. A dash (“–”) in the conditional P-value column indicates that conditional testing was not applicable because the gene was not retained as an independent signal, did not reach statistical significance, or represented the only independent gene within the locus, in which case only the primary association P-value is reported. The columns “Genes in Region (N)” and “Independent Genes in Region (n)” indicate the total number of transcriptome-wide significant genes within the locus and the number of statistically independent genes identified after conditional analysis, respectively.

**Supplementary Table 6**. Cross-ancestry transferability of European T2D-TWAS signals in the African American–ancestry (AFA) cohort. Columns summarize tissue-specific results for subcutaneous adipose, visceral omentum adipose, liver, skeletal muscle, and pancreas. “EUR-significant protein-coding genes” indicates the number of protein-coding genes identified at transcriptome-wide significance in the European-ancestry (EUR) TWAS (P < 1.919 × 10^−6^). “Genes with AFA models (N)” denotes the subset of EUR-significant genes for which predictive expression models were available in AFA and that were therefore eligible for replication analysis. Replication was evaluated by (i) nominal replication, defined as the number and proportion of genes with P < 0.05 in the AFA TWAS, and (ii) concordant effect direction, defined as the number and proportion of genes showing the same effect direction in AFA as in EUR. Proportions were calculated relative to N. One-sided binomial tests were performed to assess whether the observed proportions exceeded the null expectations of 0.05 for nominal replication and 0.5 for effect direction concordance, with corresponding binomial P values reported.

